# BinDel: detecting clinically relevant fetal genomic microdeletions using low-coverage whole-genome sequencing-based NIPT

**DOI:** 10.1101/2022.09.20.22280152

**Authors:** Priit Paluoja, Tatjana Jatsenko, Hindrek Teder, Kaarel Krjutškov, Joris Robert Vermeesch, Andres Salumets, Priit Palta

**Affiliations:** Department of Obstetrics and Gynaecology, Institute of Clinical Medicine, University of Tartu, Tartu, Estonia; Competence Centre on Health Technologies, Tartu, Estonia; Department of Human Genetics, KU Leuven, Leuven, Belgium; Division of Obstetrics and Gynecology, Department of Clinical Science, Intervention and Technology, Karolinska Institutet and Karolinska University Hospital, Stockholm, Sweden; Institute of Genomics, University of Tartu, Tartu, Estonia; Institute for Molecular Medicine Finland (FIMM), University of Helsinki, Helsinki, Finland

**Keywords:** microdeletion, BinDel, screening, DiGeorge, Angelman, Williams-Beuren, Smith-Magenis, NF1, software, NIPT

## Abstract

**Background:** Clinically pathogenic chromosomal microdeletions (MDs) cause severe fetal genetic disorders such as DiGeorge and Prader-Willi/Angelman syndromes. Motivated by the absence of reliable blood and/or ultrasound screening biomarkers for detecting microdeletion risk during the first-trimester screening, we developed and validated BinDel, a software package to evaluate the risk of clinically pathogenic microdeletions from low-coverage whole-genome-sequencing (WGS)-based NIPT data.

**Results:** We used 584 NIPT samples, including 34 clinically pre- and postnatally confirmed microdeletions, to perform a blind evaluation of the BinDel software. In a combined analysis of 34 microdeletion and 50 euploid fetal samples, BinDel correctly identified 25 samples with microdeletions in the ‘blind’ analysis. BinDel had 15 false-positive microdeletion calls, whereas the majority of them were concentrated in a few challenging regions, like NF1 microdeletion region. As a comparison, WisecondorX identified 16 correct microdeletion calls with no false-positive calls. After improving BinDel, 30 microdeletion samples were correctly determined, with a total of three false-positive microdeletion calls. Using simulated fetal microdeletions, we investigated the impact of fetal DNA fraction (FF) and microdeletion region length on BinDel’s microdeletion risk detection accuracy in 12 clinically pathogenic microdeletion regions and determined that high FF is one of the most important factors for correct MD risk detection, followed by the observation, particularly in samples with lower FF, that longer microdeletion regions exhibit higher MD risk detection sensitivity.

**Conclusions:** We confirmed BinDel feasibility for fetal microdeletion risk detection in NIPT. Remarkably, the final BinDel tool correctly identified 88.2% (30 out of 34) MD cases, opening the possibility to integrate microdeletion analysis successfully into routine NIPT protocol. Additionally, we demonstrated that high FF is one of the most important factors for correct microdeletion risk estimation and that longer microdeletion regions display higher MD calling sensitivity. This work stands as a unique contribution to prenatal microdeletion screening, exhibiting a novel software simultaneously validated with a large microdeletion sample set, positioning it as the first of its kind in the field. BinDel is available at https://github.com/seqinfo/BinDel.

## Background

Non-Invasive Prenatal genetic Testing (NIPT) is a circulating cell-free DNA (cfDNA) and sequencing-based screening technique to detect the risk of fetal aneuploidies from maternal plasma. In addition to screening for most common aneuploidies, whole-genome sequencing (WGS) NIPT assays enable the analysis of any region of the genome to infer the risk of potential (sub)chromosomal aberrations, including microdeletions (MDs). Pathogenic MDs have different incidences ranging from 1:4,000 for DiGeorge syndrome to 1:15,000-1:50,000 for MD causing Cri-du-chat syndrome (5p-) [1,2]. For example, DiGeorge syndrome, caused by the 22q11.2 heterozygous deletion, is characterised by adverse infant and childhood clinical outcomes that can include intellectual disabilities, schizophrenia, cardiovascular anomalies, hypocalcemia, palatal anomalies, obesity, hypothyroidism, hearing deficits, cholelithiasis, scoliosis, dermatologic abnormalities, etc [1,3,4].

As there are no biomarker-based screening tests for MDs for the first trimester pregnancies, and ultrasonography can only occasionally detect associated findings (e.g., enlarged nuchal translucency and/or other anatomical anomalies) associated with specific MDs, more reliable microdeletion screening methods are required [5]. Although MDs often go undiagnosed during the first-trimester combined screening, some may be identified later during the second or third-trimester, or even neonatally, if not in early childhood [6]. Fetal invasive techniques such as chorionic villus sampling and amniocentesis can confirm the presence of genetic aberrations for fetuses with abnormalities seen on ultrasound scan, but have a procedure-related risk for miscarriage [7]. An alternative approach with no risk of miscarriage would be the NIPT. However, as NIPT findings must be confirmed using invasive approaches, it is considered a screening, not a diagnostic tool. Consequently, ensuring a high positive predictive value (PPV) for microdeletion screening in NIPT is needed. There is considerable variability in positive predictive values (PPV) for the same microdeletion genomic regions among various NIPT platforms, suggesting an opportunity to improve the accuracy of microdeletion screening in NIPT [8]. This improvement in accuracy is essential to prevent unnecessary invasive procedures prompted by false positive microdeletion calls, thereby minimising the potential harm. Therefore, integrating pathogenic chromosomal microdeletion screening into routine NIPT is becoming increasingly more clinically relevant and attractive due to advancements in NIPT laboratory methods and data algorithms [9–11].

The accuracy of any NIPT protocol relies on several factors, among which the size of the fetal DNA fraction (FF) is one of the most important. FF is the proportion of cell-free DNA molecules originating from the fetus/placenta compared to the total pool of cell-free DNA molecules captured from a pregnant woman’s blood sample [12]. The higher the FF, the greater the possibility of detecting potential fetal MDs, regardless of the high maternal DNA ‘background’. The same principle applies to the MD region length [13,14], i.e., the longer the MD region, the more sequencing reads are expected to cover it, increasing the potential to detect the deficit of sequencing reads in the studied sample for a specific chromosomal region, and consequently drawing attention to a potential MD risk.

Even though there are some existing software tools that, besides detecting full chromosomal aneuploidies, can also reveal putative MDs (e.g., WisecondorX), these tools rely on heuristic anomaly detection algorithms to identify microdeletions across the entire genome and do not consider MD region-specific coordinates or characteristics [15,16]. However, in the clinical context, clinically relevant pathogenic MDs typically have well-defined genomic coordinates. Thus, concentrating solely on such a predefined list of genomic coordinates of clinically significant MD regions will prevent recognising the variants of uncertain significance (VUS), avoiding using invasive pregnancy follow-ups in VUS cases.

Here, we present BinDel, a novel software tool developed to infer a priori chosen clinically relevant MD risk estimates from low-coverage WGS-based NIPT data (**Fig. 1, Suppl. Table S1**). In order to assess the ability of BinDel to detect MD risks under various conditions, we investigated the impact of FF and microdeletion region length on MD detection accuracy using NIPT samples with computationally simulated fetal microdeletions. Finally, we validated BinDel ‘blindly’ using 50 euploid and 34 clinically confirmed samples that included MDs from several specific regions, encompassing neurofibromatosis type 1 (NF1), DiGeorge, Williams-Beuren, Smith-Magenis, and Prader-Willi/Angelman syndrome associated MD regions (**Suppl. Table S2**).

**Fig. 1.**
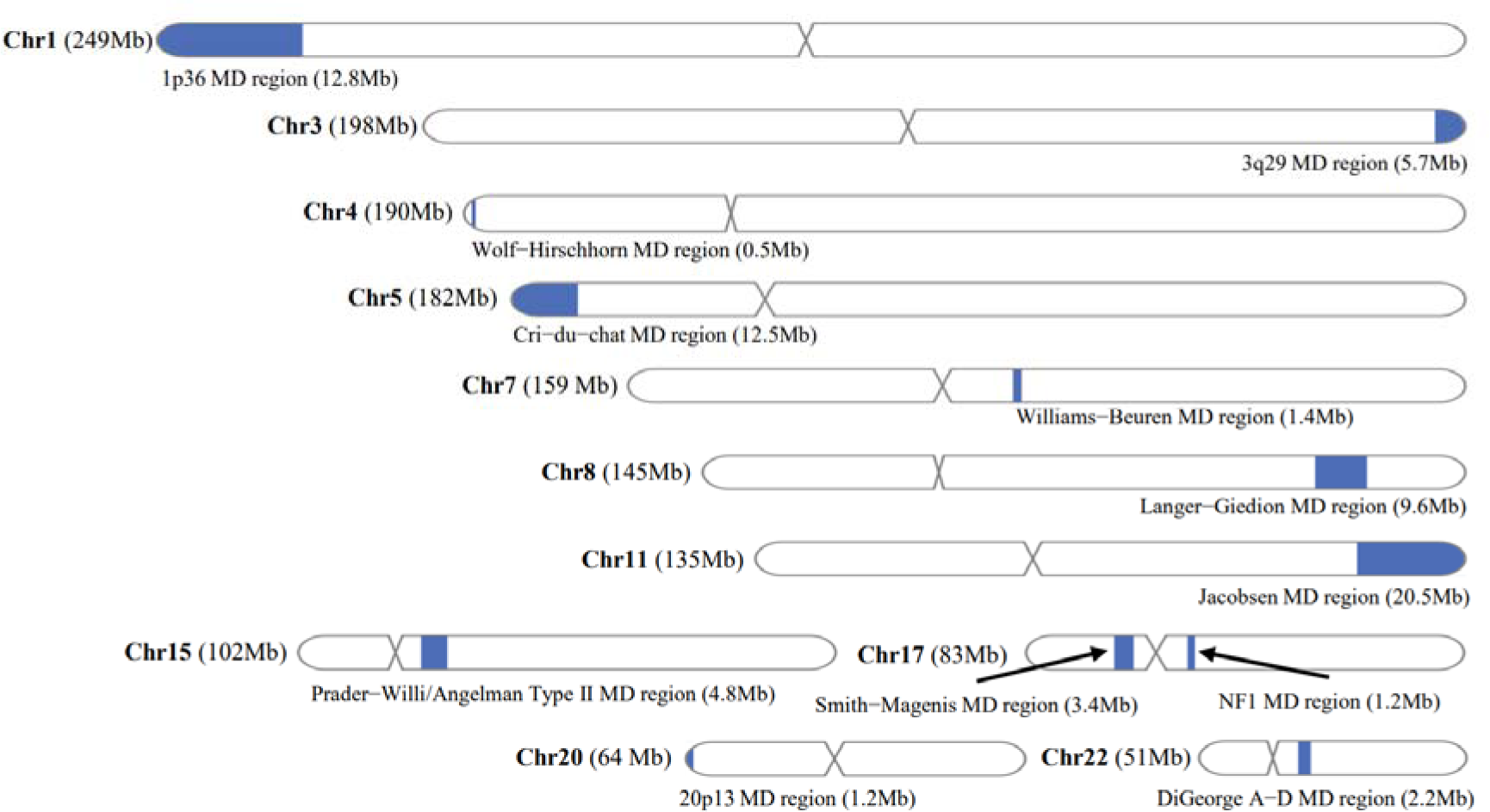
A priori chosen clinically relevant microdeletions (MD) and their locations on the genome. Microdeletion regions are indicated with blue sections on the chromosomes.

## Materials and Methods

### Ethics statement

This study was performed with the written informed consent from the participants and with the approval of the Research Ethics Committee of the University of Tartu (#352/M-12) and the Ethics Committee Research UZ / KU Leuven (S66817).

### BinDel algorithm

BinDel is a novel microdeletion detection R package to infer MD risk from low-coverage WGS NIPT data. One significant strength of BinDel is its targeted approach to microdeletion risk screening in WGS-based NIPT, focusing on predefined configurable genomic coordinates (e.g., DiGeorge MD regions A-B or A-D) rather than serving as an exploratory heuristic algorithm throughout the entire genome. Unlike exploratory algorithms, which may not provide an MD risk estimate if they interpret the MD as noise, BinDel’s coordinate-based approach guarantees that the target MD region always receives an MD risk estimate. In the clinical context, pathogenic MDs causing genetic syndromes, such as DiGeorge syndrome, typically have well-defined genomic coordinates, making the targeted approach highly suitable for NIPT.

From an algorithmic perspective, the inference of MD risk begins with the division of aligned sequencing reads into a priori selected and named (certain bins denote MD regions, others non-MD regions) genomic bins, followed by a series of normalisation steps. Initially, the genomic bins undergo GC% correction, and subsequently, they are further normalised based on the total sequencing read count and bin lengths [17]. Next, the normalised values are divided by principal component analysis (PCA) dimensionality-reduced versions [18].

The PCA normalisation method in BinDel involves transforming the input data by projecting it onto a reduced-dimensional space using PCA. Initially, the input data frame contains genomic bins as columns (e.g., chr1:100000, chr1:200000, etc.), with rows representing samples and values indicating the normalised read counts of the bins.

The prcomp function in R is employed to perform PCA on the data, retaining the first N principal components. Subsequently, the original data is reconstructed with reduced dimensionality by multiplying the principal component matrix with the transposed rotation matrix, considering only the first N components.

To revert the reconstructed data back to its original scale, it is adjusted with a negative mean obtained during the centring process before PCA. Finally, the pre-PCA-transformed data is divided by reconstructed data. This analysis is applied first to the euploid fetus reference group, and then, using the parameters obtained, the same analysis is applied to the sample under study. Using PCA normalisation, BinDel reduces the impact of irrelevant factors that may interfere with MD risk detection.

Following PCA-normalisation using a reference set, BinDel calculates both a Z-score and a ‘normalised’ Z-score for each genomic bin using the euploid fetus reference set. The ‘normalised’ Z-score accounts for the total number of bins that exceed the mean normalised read count in the target genomic region, and both Z-scores are divided by the square root of the number of bins in the target region before being summed per target region in two separate features.

Next, based on the aggregated features, BinDel calculates the Mahalanobis distance for each MD region from the euploid fetus reference group. This distance measure helps identify outliers, potentially indicating MD risk, by comparing the distance of each region relative to the centroid of the euploid reference group target genomic regions.

To ensure uniform interpretation and comparability between different genomic regions, the Mahalanobis distances are transformed into chi-squared distribution p-values and subsequently −log10 transformed. This step restricts the microdeletion risk output to a range between 0% and 100%, allowing for consistent understanding and comparison of MD risk levels independent of the length or variability of the regions.

BinDel offers various metrics in its output, distinguished by the prefixes or suffixes ‘conservative’ and ‘greedy’. The primary difference lies in whether the metrics utilise a single aggregated and ‘normalised’ Z-score (‘conservative’) or also consider aggregated but ‘non-normalised’ Z-scores (‘greedy’).

BinDel is available from GitHub (https://github.com/seqinfo/BinDel/).

### Sample pre-processing, microdeletion high-risk calling criteria, genomic bin sizes, and a priori chosen target microdeletion region coordinates

Each sequenced sample was aligned against human reference assembly GRCh38 using BWA-MEM, sorted, and the reads originating from the same fragment of DNA were marked as duplicates using MarkDuplicatesSpark [19,20].

BinDel output ‘conservative probability’ was used to call MD risk probability. To determine the high MD risk call from the BinDel output, we established specific criteria based on the MD risk probability for shorter regions such as NF1 and Williams-Beuren MD region: a minimum probability of 50% with PCA99% (which captures 99% of the cumulative variance). An 80% cut-off was used with PCA95% for longer target microdeletion regions.

In order to maintain an adequate number of bins per MD region and reduce fluctuations caused by random sequencing read placement effects and varying sequencing read placement patterns between different microdeletion regions, after analysing different bin sizes, 300kb bin size was used [21]. However, in shorter regions, where 300kb led to too few bins, 100kb for Wolf-Hirschhorn and DiGeorge B/C-D MD and 200kb for NF1 MD were used (**Suppl. Table S1**). The coordinates for the target microdeletion regions were mainly derived from the DECIPHER, ClinGen and OMIM databases [22–24] (**Suppl. Table S1**).

### Simulation methodology for assessing microdeletion risk detection sensitivity and specificity

#### Studied samples

We used 300 samples reported previously as euploid fetus pregnancies by NIPTIFY screening test and postnatal evaluation at Competence Centre on Health Technology (CCHT, Tartu, Estonia). Samples were processed similarly to previously published guidelines from Katholieke Universiteit Leuven, with modifications [25]. Peripheral blood samples were collected in cell-free DNA BCT tubes (Streck, USA), and plasma was separated with standard dual centrifugation. Cell-free DNA was extracted from 3 ml plasma using MagMAX Cell-Free DNA Isolation Kit (ThermoFisher Scientific). Whole-genome libraries were prepared using the FOCUS (Fragmented DNA Compact Sequencing Assay) NIPT protocol at CCHT with 12 cycles for the final PCR enrichment step. Following library quantification, 36 samples were pooled equimolarly, and the quality and quantity of the pool were assessed on Agilent 2200 TapeStation (Agilent Technologies, USA). WGS was performed on the NextSeq 550 instrument (Illumina Inc.) 85 bp single-end with an average coverage of 0.32×. Out of these 300 samples, 200 were utilised as a reference set for BinDel and the remaining 100 were employed for simulating fetal microdeletions. The read count ranged from 8M to 18.5M reads per sample (RPS), with a median of 12.7M RPS.

#### Simulation process and data analysis

We performed the following simulation to assess the FF, MD region length, and the effect of the number of PCA components on MD risk detection sensitivity and specificity. For each a priori chosen MD region, we iterated over 100 normal samples (**Suppl. Table S1**). For each sample within the nested loop, we further iterated over a set of target FF, including FF of 0%, indicating a sample retaining all reads later used as negative MD control for MD risk detection specificity calculations. Inside the innermost loop, the simulation of heterozygous fetal MD in the target region was performed using the GATK (Genome Analysis Toolkit [19]).

First, the reads within the MD region were extracted from the input Binary Alignment Map (BAM) file, downsampled to mimic deletion with target fetal fraction (for example, with a target FF of 20%, 90% of reads are retained), and output to a temporary BAM file. Next, the reads outside the MD region were extracted from the input BAM file to another temporary BAM file. Finally, the two temporary BAM files were merged into a single BAM, resulting in a BAM file with simulated heterozygous fetal MD. The final BAM file was analysed with BinDel using 129 and 183 PCA components. Specifically, 129 and 183 were chosen as they correspond to capturing 95% and 99% of the cumulative variance within the BinDel reference set, respectively.

Upon completing the nested loop iterations, we calculated the sensitivity and specificity considering different target FFs, the number of PCA components and target MD regions.

### BinDel ‘blind’ validation

We had 200 known fetal euploid samples for reference set needed by BinDel and WisecondorX (v1.2.4), and 84 ‘blind’ samples containing 50 euploid and 34 samples having clinically confirmed MDs in DiGeorge, Prader-Willi/Angelman, Smith-Magenis, Williams-Beuren and NF1 MD regions. These 34 samples originated from 22 unique patients, from which some cfDNAs were sequenced 2-3 times in different libraries (**Suppl. Table S2**). Of 22 unique patients, 3 had co-occurring maternal and fetal deletions, one had solely maternal MD, and the remaining 18 were fetal-only MDs (**Suppl. Table S2**). The single-end FASTQ files were aligned and processed, as mentioned before. The ‘blind’ sample set FF values were between 3% and 16.5% with a median of 8.1%, and the read count ranged from 7.5 to 39.2M RPS, with a median of 13.2M RPS.

We applied both BinDel and WisecondorX (default parameters) on 84 samples, reported the high-risk MD calls to Katholieke Universiteit Leuven, and received sample states (euploid or MD) for each sample analysed. We counted the number of false-positive, false-negative and true-positive MD risk calls.

### Improving BinDel microdeletion risk detection

After ‘blind’ validation, we investigated further, departing from the previous ‘blind’ approach, to determine if it is feasible to establish an improved high-risk MD cut-off threshold and whether the development of a novel method of applying secondary per-MD-region PCA normalisation after the initial PCA normalisation could enhance microdeletion detection sensitivity and specificity. The regional cumulative PCA normalisation method is a PCA normalisation applied separately to each specific target region, taking into account the cumulative variance within that region following the initial ‘general’ PCA normalisation. To consider both the number of true-positive MD calls and false-positive MD calls, we assessed the F-measure, a metric reflecting the harmonic mean of positive predictive value and sensitivity (where higher values indicate superior performance).

First, we investigated across 1-100% decision points for microdeletion high-risk cut-off using PCA95% if we could find a single cut-off threshold over target regions providing higher F-measure than F-measure achieved from the ‘blind’ analysis.

Secondly, we investigated if we could find a combination of cumulative PCA and regional PCA and cut-off threshold per target region that would achieve the highest F-measure. We analysed the validation sample set with combinations of PCA95%, PCA99%, cut-offs 1%-100% and cumulative regional PCA of 5%, 10%, 15%, 25% and 50% (only for PCA95%). In instances where multiple combinations yielded identical maximum F-measures, preference was given to those with lower PCA and regional PCA cumulative values and cut-off thresholds.

## Results

In this study, we present BinDel, a novel software package to infer MD risk in predefined regions of interest from low-coverage WGS NIPT data. We developed BinDel by using simulated MD NIPT data and validated it on clinically confirmed MD NIPT samples.

### Fetal fraction and region length effect on BinDel performance accuracy

First, we investigated the impact of FF and microdeletion region length on MD detection accuracy using NIPT samples with computationally simulated fetal microdeletions in 1p36, 20p13, 3q29, Cri-du-chat, DiGeorge, Jacobsen, Langer-Giedion, NF1, Prader-Willi/Angelman, Smith-Magenis, Williams-Beuren and Wolf-Hirschhorn MD regions (**Suppl. Table S1**).

#### Fetal DNA fraction

As expected, we observed that fetal DNA fraction had a significant effect on MD detection accuracy. Higher FF considerably increased the MD detection sensitivity (**Fig. 2A-D, Suppl. Fig. S1**). However, this pattern varied across different MD regions. Specifically, when comparing the Williams-Beuren or DiGeorge A-B MD region to the NF1 MD region, the MD risk-calling sensitivity in the NF1 MD region was higher across all FF levels despite having a similar MD length (**Fig. 2A-C)**.

**Fig. 2.**
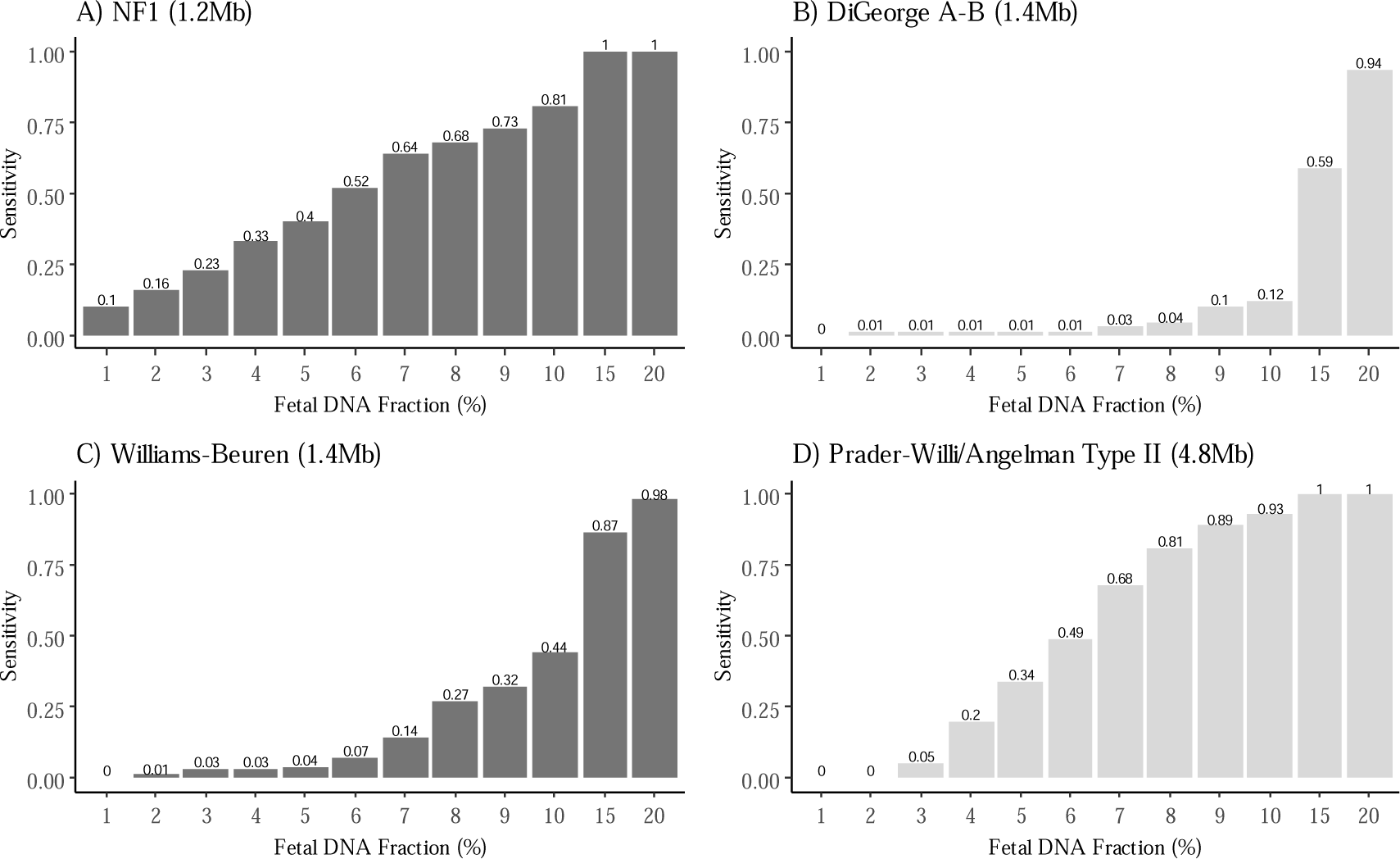
The effect of fetal DNA fraction on BinDel microdeletion risk detection sensitivity. Sensitivity estimates are indicated by grey and dark grey bars, corresponding to PCA95% and PCA99%, respectively. Sensitivity is calculated for NF1 (A), DiGeorge A-B (B), Williams-Beuren (C) and Prader-Willi/Angelman (D) associated microdeletion regions.

We also observed the impact of microdeletion region length on MD calling accuracy. Our analysis revealed a reoccurring trend whereby longer MD regions demonstrated significantly higher MD risk detection sensitivity, particularly evident in the case of samples with lower FF (relates to **Fig. 2D, Fig. 3A-B**). With some exceptions, MD risk detection in shorter MD regions proved challenging even at higher FF levels (**Fig. 3D**). For example, the relatively short NF1 region (1.2Mb) exhibited sensitivity somewhat comparable to the longer Prader-Willi/Angelman MD region (4.8Mb), even at lower FF levels, highlighting differences in MD risk calling in different well-established MD regions (**Fig. 3A-D**).

**Fig. 3.**
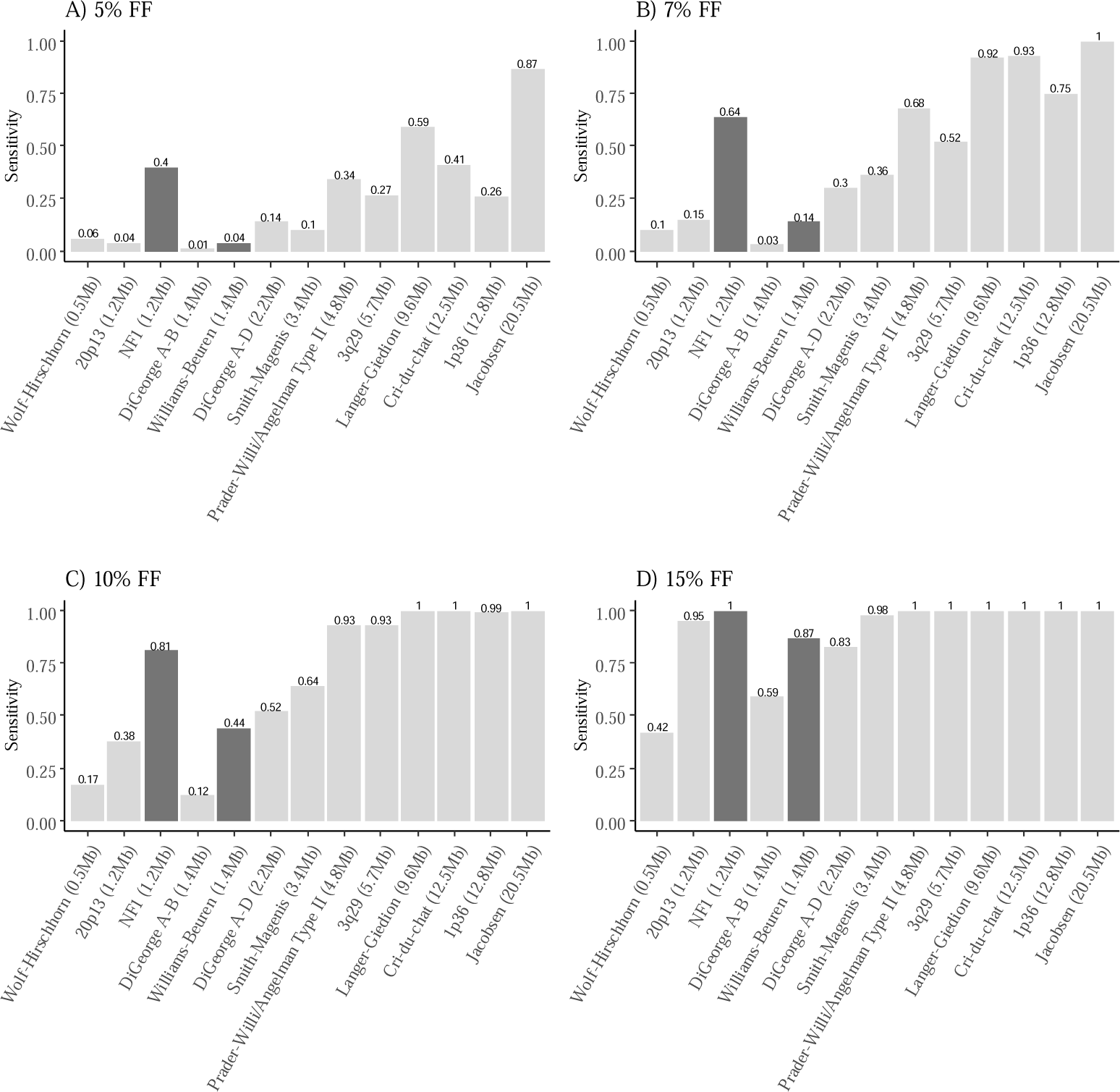
Microdeletion associated region length effect on the BinDel sensitivity of detecting microdeletion risk. Fetal DNA fractions of 5% (A), 7% (B), 10% (C) and 15% (D) were used for *in silico* simulated microdeletions. Sensitivity estimates are indicated by grey and dark grey bars, denoting PCA95% and PCA99%, respectively.

#### Improving the accuracy of MD risk detection in short microdeletion regions

To increase the MD risk detection sensitivity in short MD regions, we examined the effect of sequencing read data normalisation by considering the number of PCA components used by BinDel in the normalisation step. We used a set of 100 euploid fetus samples to compute specificity, while for sensitivity calculations, we simulated fetal microdeletions as outlined in the Materials and Methods section. We evaluated two normalisation settings, PCA95% and PCA99% (see **Methods**). We observed that using PCA99% for normalisation increased MD detection sensitivity, particularly in short MD regions like NF1 (1.2Mb), where MD detection sensitivity increased 2.5-fold from 0.16 to 0.4 at an FF of 5% (**Fig. 4A1**). However, this was not the case in all MD regions. For example, in the case of the Williams-Beuren MD region, the overall sensitivity and specificity, when using PCA99%, were either equal or slightly lower as compared to the lower PCA95% setting (**Fig. 4B1, B2**). It is also important to note that using PCA99% did not only result in higher MD risk detection sensitivity but also resulted in an increase in false-positive MD risk calls in certain regions, such as DiGeorge A-D, once more highlighting the unique characteristics of each MD region (**Fig. 4C2, Table 1**). Within microdeletion regions like DiGeorge, encompassing multiple potential intra-region deletion variants (e.g., A-B or A-D), maximising sensitivity and specificity justifies the rationale for BinDel to screen and consider all deletion variants.

**Fig. 4.**
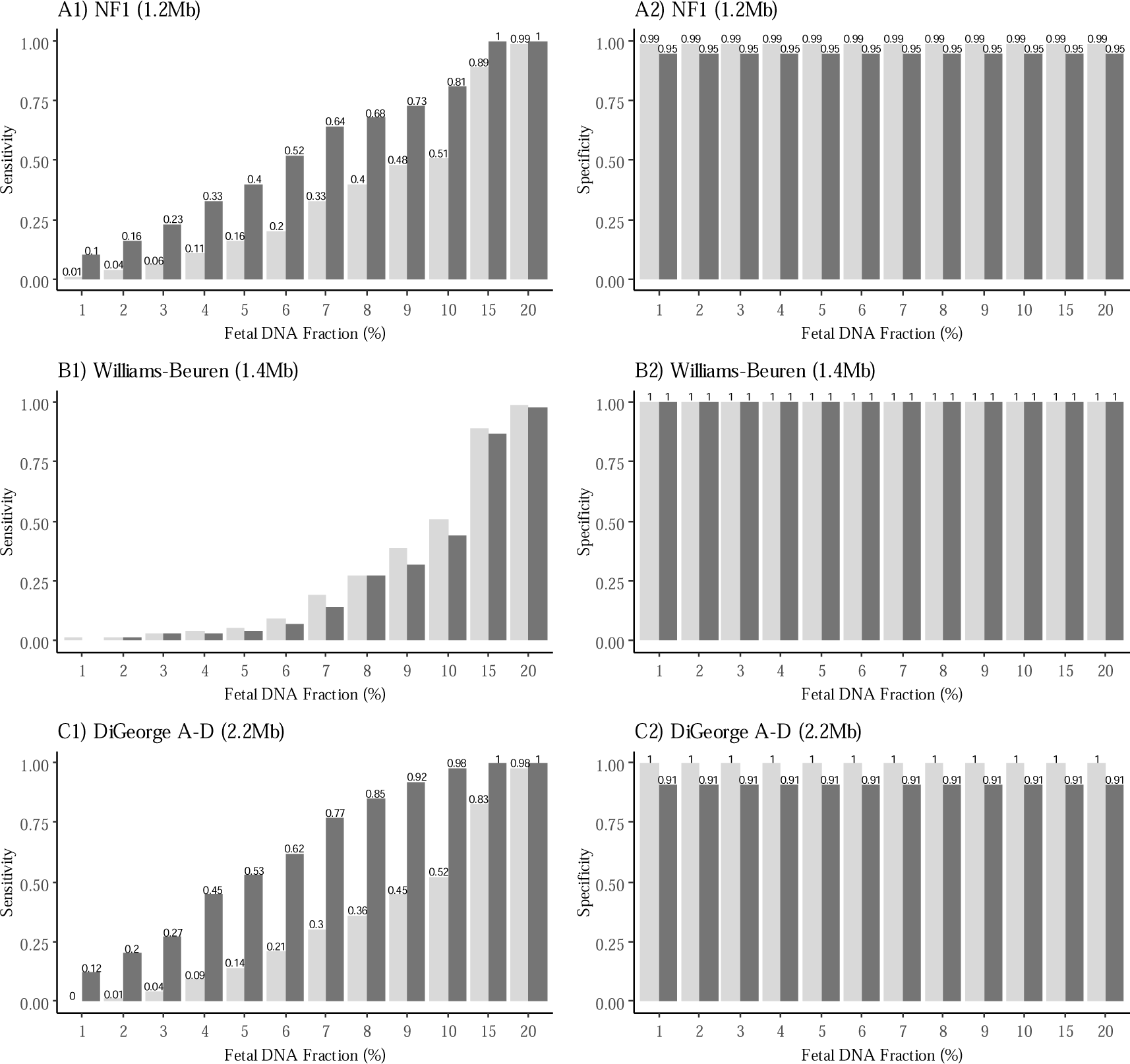
BinDel sensitivity and specificity with *in silico* simulated microdeletions using different numbers of PCA components. Sensitivity estimates are indicated by grey and dark grey bars, denoting PCA95% and PCA99%, respectively. The analysis focuses on three MD regions: NF1 (A1, A2), Williams-Beuren (B1, B2), and DiGeorge A-D (C1, C2) associated microdeletion regions.

**Table 1.** BinDel false-positive microdeletion risk calls. False-positive microdeletion risk calls in different microdeletion regions across a cohort of 100 samples with euploid fetuses (placenta) determined by NIPT.

| Microdeletion region | Percentage of false-positive calls: PCA95% | Percentage of false-positive calls: PCA99% |
| --- | --- | --- |
| Wolf-Hirschhorn (0.5Mb) | 0 | 4 |
| 20p13del (1.2Mb) | 0 | 2 |
| NF1 (1.2Mb) | 1 | 5 |
| DiGeorge A-B (1.4Mb) | 0 | 4 |
| Williams-Beuren (1.4Mb) | 0 | 0 |
| DiGeorge A-D (2.2Mb) | 0 | 9 |
| Smith-Magenis (3.4Mb) | 1 | 1 |
| Prader-Willi/Angelman (4.8Mb) | 0 | 2 |
| 3q29 (5.7Mb) | 0 | 4 |
| Langer-Giedion (9.6Mb) | 0 | 5 |
| Cri-du-chat (12.5Mb) | 0 | 1 |
| 1p36 (12.8Mb) | 0 | 4 |
| Jacobsen (20.5Mb) | 0 | 1 |

### Evaluation of BinDel with clinically validated microdeletion samples

In order to estimate BinDel accuracy, we conducted a ‘blind’ analysis using 84 samples containing 34 clinically confirmed MD and 50 confirmed euploid NIPT samples. Initially, BinDel correctly detected 73.5% (25 out of 34) clinically validated samples as high risk (**Table 2, Suppl. Table S3**). When considering samples only with fetal microdeletions, excluding co-occurring maternal and fetal or one solely maternal MDs, BinDel initially correctly detected 69% (19 out of 28) of fetal MD samples (**Table 2, Suppl. Table S3**). For example, BinDel detected 12 out of 17 (71%) samples with DiGeorge fetal microdeletion as high-risk. On the other hand, BinDel failed to correctly identify any of the three samples with Williams-Beuren microdeletion as high risk. Emphasising the importance of FF, we observed that the average FF of missed fetal MD samples was lower (8.9%) as compared to those where fetal MDs were correctly called (10.4%). In the DiGeorge MD region, BinDel had no false-positive MD calls. A noteworthy exception was NF1 MD region, where we observed a notable occurrence of false-positive high-risk calls. In parallel, we also inferred MD calls with WisecondorX for the same samples (**Table 2, Suppl. Table S3**). WisecondorX showed similar sensitivity in the Smith-Magenis and Williams-Beuren MD regions. In other MD regions, there were differences in MD risk detection sensitivity (**Table 2)**. On the other hand, WisecondorX had no false-positive MD calls in any of the five MD regions analysed for these samples.

**Table 2.** BinDel and WisecondorX accuracy in ‘blind’ analysis. The number of true positive (TP), false positive (FP), false negative (FN), and positive predictive values (PPV) of microdeletion high-risk calls of BinDel and WisecondorX software packages on clinically confirmed 34 microdeletion and 50 euploid NIPT samples.

| Microdeletion region, origin and<br>number of samples (n) | BinDel |  |  |  | WisecondorX |  |  |  |
| --- | --- | --- | --- | --- | --- | --- | --- | --- |
|  | TP | FP | FN | PPV | TP | FP | FN | PPV |
| DiGeorge, fetal, n = 17 | 12 | 0 | 5 | 1 | 4 | 0 | 13 | 1 |
| DiGeorge, maternal and fetal, n = 4 | 4 | 0 | 0 | 1 | 4 | 0 | 0 | 1 |
| DiGeorge, maternal, n = 1 | 1 | 0 | 0 | 1 | 1 | 0 | 0 | 1 |
| Prader-Willi/Angelman, fetal, n = 5 | 5 | 2 | 0 | 0.71 | 4 | 0 | 1 | 1 |
| Smith-Magenis, fetal, n = 3 | 2 | 0 | 1 | 1 | 2 | 0 | 1 | 1 |
| Williams-Beuren, fetal, n = 3 | 0 | 3 | 3 | 0 | 0 | 0 | 3 | 0 |
| NF1, maternal and fetal, n = 1 | 1 | 10 | 0 | 0.09 | 1 | 0 | 0 | 1 |

### Improving microdeletion risk detection

Considering the previously described ‘blind’ calling procedure, it was apparent that MD detection accuracy varies considerably across different genomic MD regions, as also seen in the simulated data presented earlier (**Table 2, Fig. 2)**. Hence, we systematically tested if we could improve microdeletion risk detection. First, we determined that increasing (see **Methods**) the MD high-risk cut-off for all MD regions to 90% and using only PCA95% reduces the occurrence of false-positive microdeletion calls from 15 to 1 while preserving the number of true-positive microdeletion calls (**Table 3, Suppl. Fig. S2)**.

**Table 3.** Microdeletion cut-off threshold effect on BinDel accuracy. The number of true-positive (TP), false-positive (FP), false-negative (FN), and positive predictive values (PPV) of microdeletion high-risk calls by BinDel with a PCA95% on different microdeletion high-risk cut-off thresholds.

| Microdeletion high-risk cut-off threshold (%) | TP | FP | FN | PPV |
| --- | --- | --- | --- | --- |
| 10 | 27 | 19 | 7 | 0.59 |
| 20 | 26 | 9 | 8 | 0.74 |
| 30 | 25 | 5 | 9 | 0.83 |
| 40 | 25 | 5 | 9 | 0.83 |
| 50 | 25 | 4 | 9 | 0.86 |
| 60 | 25 | 3 | 9 | 0.89 |
| 70 | 25 | 3 | 9 | 0.89 |
| 80 | 25 | 3 | 9 | 0.89 |
| 90 | 25 | 1 | 9 | 0.96 |

Secondly, we considered per MD region factors beyond region length, such as regional patterns in read count distribution. To overcome the latter challenge, we added an additional regional PCA normalisation method (see **Methods, Suppl. Fig. S3**), which was applied systematically to all MD regions combined with regionally different high-risk cut-off thresholds. This approach detected 30 out of 34 (88.2%) MD samples with only three false positive MD calls, marking a 14.7% increase (from 73.5% to 88.2%) in detection sensitivity compared to the ‘blind’ validation analysis (**Table 2, Table 4, Suppl. Table S3)**. Especially noteworthy is the Williams-Beuren MD region, where BinDel identified 2 out of 3 true-positive samples with the novel regional PCA normalisation compared to the previous no-call results (**Table 4, Suppl. Table S3)**. Again, we observed that the average FF for four samples not correctly identified as MD samples was 8% compared to 10% for 30 correctly identified MD samples, confirming the previously observed FF effect on MD detection accuracy.

**Table 4.** Region specific approach effect on microdeletion screening accuracy. The number of true-positive (TP), false-positive (FP) and positive predictive values (PPV) of microdeletion calls by BinDel with a region-specific approach. The number of true-positive high-risk microdeletion calls increased compared to the ‘blind’ analysis in DiGeorge, Smith-Magenis and Williams-Beuren microdeletion regions. False-positive microdeletion calls are reduced in Prader-Willi/Angelman, NF1 and Williams-Beuren microdeletion regions.

| **Microdeletion, origin and number of samples (n)** | **Microdeletion high-risk cut-off threshold (%)** | **Cumulative PCA (%)** | **Cumulative regional PCA (%)** | **TP** | **FP** | **PPV** |
| --- | --- | --- | --- | --- | --- | --- |
| DiGeorge<br>fetal, fetal and maternal,<br>maternal<br>n = 19 | 9 | 95 | Not used | 19 | 0 | 1 |
| Prader-Willi/Angelman<br>fetal<br>n = 5 | 87 | 95 | Not used | 5 | 1 | 0.8 |
| Smith-Magenis<br>fetal<br>n = 3 | 4 | 95 | Not used | 3 | 0 | 1 |
| Williams-Beuren<br>fetal<br>n = 3 | 10 | 95 | 50 | 2 | 2 | 0.5 |
| NF1<br>maternal and fetal<br>n = 1 | 87 | 95 | Not used | 1 | 0 | 1 |

## Discussion

Clinically pathogenic fetal microdeletions are chromosomal abnormalities that can have a profound impact on fetal and child development and health. Here, we present BinDel, a novel MD detection software to infer microdeletion risk from low-coverage WGS-based NIPT data. We performed comprehensive analyses with simulated fetal microdeletion samples to evaluate the influence of fetal DNA fraction (FF) and MD region length on BinDel MD risk detection accuracy. Subsequently, a ‘blind’ validation study was carried out to validate BinDel’s sensitivity and specificity in detecting microdeletion risk. For this, we used 34 NIPT samples from confirmed MD cases, including the samples with NF1, DiGeorge, Williams-Beuren, Smith-Magenis, and Prader-Willi/Angelman MD regions, with an additional 50 euploid fetal NIPT samples (**Suppl. Table S2**). After ‘blind’ MD calling, we were able to improve MD risk detection even further, by adding a novel regional PCA normalisation method. Using this updated version, a remarkable proportion (88.2%, 30 out of 34) of MD samples were correctly determined (**Table 4**).

Besides sequencing coverage, we confirmed that FF is one of the most important factors for correct MD detection (**Fig. 2A-D, Suppl. Fig. S1A-I**) [14]. MDs of 4.8Mb and longer were determined with 100% sensitivity from FF of 15% (**Fig. 3**). Therefore, as also demonstrated by Welker *et al*., increasing the FF through laboratory enrichment procedures would potentially be a key component to increase the MD risk detection [26]. We also observed that the MD samples that were not correctly identified had slightly lower average FF than the correctly detected MD samples. Moreover, three Williams-Beuren MD samples not detected by BinDel in the ‘blind’ validation sub-study had relatively low FF values of 6.38%, 7.43% and 8.86%, concurring with those observed in FF analysis performed with simulated MD data (**Fig. 2C**).

In the ‘blind’ validation sub-study, BinDel correctly identified 73.5% (25 out of 34) clinically confirmed MD samples of fetal, co-occurring maternal and fetal, and solely maternal origin, or 69% (19 out of 28) of solely fetal MD samples (**Table 2, Suppl. Table S3)**. We also tested WisecondorX, a universal WGS copy number detection tool not intended only for NIPT, but for broader genomic applications [15]. WisecondorX correctly identified 16 samples with microdeletions, from which 10 were fetal only (**Table 2, Suppl. Table S3**). Notably, WisecondorX had no FP MD calls in any of the target microdeletion regions, whereas BinDel had two FP MD calls in Prader-Willi/Angelman, three in Williams-Beuren and 10 in NF1 MD region, and no FP MD calls in DiGeorge and Smith-Magenis MD regions. As expected, both tools detected all samples with maternal and fetal deletion or maternal deletion only (**Table 2**).

Departing from the ‘blind’ validation and following the development of a novel regional PCA normalisation (**Suppl. Fig. S3A-C**) with a region-specific MD cut-off threshold, BinDel successfully identified 30 out of the 34 samples, i.e., 88.2% of the test MD samples (**Table 4**). With a region-specific approach, the BinDel FP MD call count was reduced from fifteen to three. The most challenging microdeletion region for BinDel was the Williams-Beuren MD region, where a low detection rate was also witnessed by Tian *et al.* [27]. BinDel’s capacity to detect samples with microdeletions in the Williams-Beuren MD region was only realised after undergoing region-specific parameter search, as evident in **Table 4**. In cases where a region-specific approach is not feasible, we advocate the adoption of PCA95% in conjunction with a microdeletion cut-off threshold set at 90%. However, when circumstances permit, specifically, when NIPT samples featuring MDs are available for calibration, we strongly recommend adopting a region-specific normalisation approach. This calibration should encompass key parameters such as PCA, regional PCA, and the cut-off threshold, tailored to align with the application’s specific requirements, ensuring a balance between sensitivity and specificity that aligns with the intended analysis purposes.

Screening for MDs poses inherent challenges. For example, microdeletion risk identified from WGS NIPT may stem from fetal, co-occurring maternal and fetal, or exclusively maternal origin, as observed from ‘blind’ analysis (**Suppl. Table S2**). Next, MD regions are often in the vicinity of low-copy repeat or homologous sequences, which, in the case of short read sequencing methods, do not facilitate unambiguously unique sequencing read mapping [28]. For instance, in the case of the DiGeorge MD region, Campbell *et al.* have demonstrated that while the majority of patients (84%) exhibit the standard A-D deletion, a significant proportion of patients possess alternative deletions, such as A-B (5%) or B-D (4%) [29]. In our BinDel validation group, we also encountered samples with DiGeorge MD region deletions of different types, namely A-B or A-D deletions (**Suppl. Table S2**). It has been observed that NF1 pseudogenes can occur on multiple chromosomes, including 15q11.2 and 22q11, which correspond to the Prader-Willi/Angelman and DiGeorge MD region, respectively [30,31]. The possible presence of NF1 pseudogenes in these regions could potentially explain why BinDel called FP NF1 MD high-risk (PCA99%, MD risk call threshold 50%, see **Methods**) for three samples with MD in Prader-Willi/Angelman and four in the DiGeorge MD region (**Table 2**) [30,31]. Additionally, the Prader-Willi/Angelman MD region deletions also have different subtypes, such as longer type I or shorter type II, the latter also found in our validation sample group [32]. Moreover, our observations indicated that even among short MD regions of similar size, the sensitivity of microdeletion risk detection varied depending on the specific MD region. This was exemplified by the sensitivity simulations of the NF1 (1.2Mb) and Williams-Buren MD (1.4Mb) regions, where the shorter NF1 MD region exhibited higher MD risk detection sensitivity (**Fig. 4A1, B1**).

In our study, we used simulated samples to calibrate the number of PCA components to use in the analysis. However, the simulations may not fully capture the complexity of actual fetal microdeletions, e.g., all possible microdeletion breakpoints. Consequently, the chosen more sensitive BinDel parameters based on the simulations for the Williams-Beuren and NF1 region resulted in the ‘blind’ validation sub-study a considerable number of false-positive risk calls in these two regions (**Table 1, Table 2**). A key distinguishing feature of BinDel is the focusing on pre-defined configurable genomic coordinates rather than serving as an exploratory algorithm throughout the entire genome. This sets BinDel apart from algorithms that rely on heuristic approaches to identify microdeletions across the entire genome. By incorporating known coordinates, BinDel eliminates the need for the determination of microdeletion start and end positions, thereby preventing potential dilution of the microdeletion signal strength resulting from boundary estimation. However, BinDel is limited in detecting microdeletion risks from regions with undefined or suboptimal coordinates. This limitation becomes apparent when a priori chosen MD region coordinates do not account for different forms of specific MDs, such as the various A-D, A-B, A-C, etc., forms in the DiGeorge MD region. If the microdeletion region is thoroughly studied, as may be the case with pathogenic microdeletions, the risk of defining suboptimal coordinates can be effectively mitigated.

## Conclusions

We developed a software tool for MD detection in WGS-based NIPT and evaluated it with a number of positive and negative control samples whilst also considering microdeletion region characteristics. In the ‘blind’ validation study, BinDel correctly identified 73.5% of clinically confirmed MD samples. Using a region-specific normalisation approach, BinDel detected 88.2% of MD samples with a total of three false positive MD calls. Furthermore, we also demonstrated that high FF is one of the most important factors for correct MD risk detection, followed by the observation that longer microdeletion regions display higher microdeletion risk detection sensitivity, more evident in samples with lower FF.

To our knowledge, this study represents one of the largest and most comprehensive efforts to date, showcasing the effectiveness of the BinDel in identifying fetal MD cases through WGS-based NIPT testing. These promising findings contribute substantially to the ongoing dispute regarding the inclusion of MD syndrome screening within NIPT protocols. The developed pipeline and analysis outcomes underscore the feasibility of detecting MD syndromes through NIPT, thereby broadening the spectrum of fetal genetic conditions detectable through this method during the first trimester. Early MD screening is crucial as these microdeletions significantly impact the child’s health.

## Availability of data and materials

BinDel source code is publicly available at GitHub (https://github.com/seqinfo/BinDel/) under the Attribution-NonCommercial-ShareAlike 4.0 International (CC BY-NC-SA 4.0) license.

The sequencing data used for this study’s simulation process and data analysis are not openly available due to sensitivity and are available from the CCHT Data Access Committee upon reasonable request. Data is located in controlled access data storage at the European Genome-phenome Archive (EGA) under accession code EGAD00001009512.

The ‘blind’ analysis sequencing data supporting this study’s findings are not openly available due to reasons of sensitivity and are available from the corresponding author upon reasonable request. Data is located in controlled access data storage at Katholieke Universiteit Leuven.

## Data Availability

BinDel source code is publicly available at GitHub (https://github.com/seqinfo/BinDel/) under the Attribution-NonCommercial-ShareAlike 4.0 International (CC BY-NC-SA 4.0) license.
The sequencing data used for this study's simulation process and data analysis are not openly available due to sensitivity and are available from the CCHT Data Access Committee upon reasonable request. Data is located in controlled access data storage at the European Genome-phenome Archive (EGA) under accession code EGAD00001009512.
The 'blind' analysis sequencing data supporting this study's findings are not openly available due to reasons of sensitivity and are available from the corresponding author upon reasonable request. Data is located in controlled access data storage at Katholieke Universiteit Leuven.

## Acknowledgements

The authors would like to thank Signe Mölder, Kate Elizabeth Stanley, and Ilse Parijs for their support and assistance.

## Supplementary Information

**Suppl. Fig. S1.**
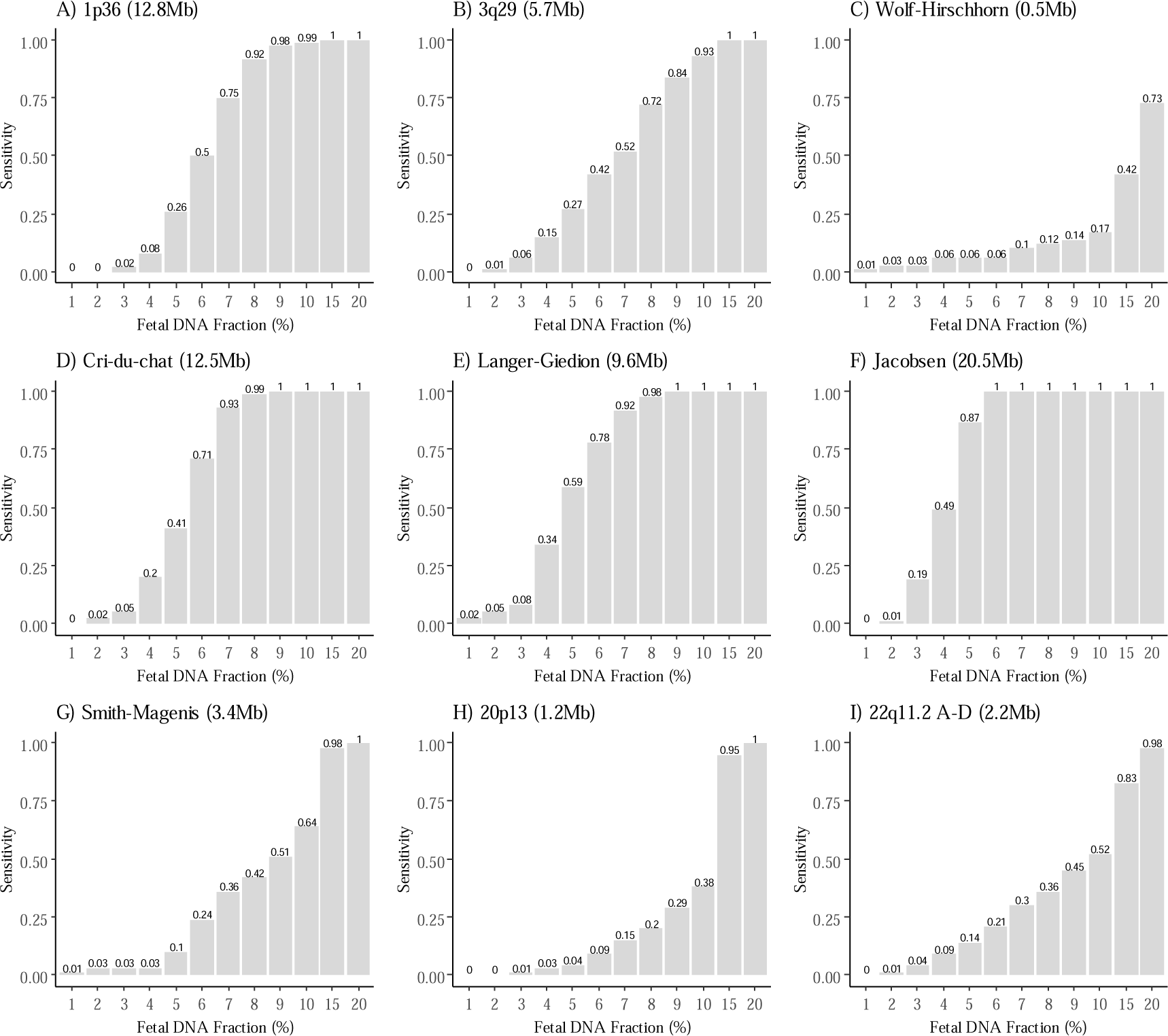
The effect of fetal DNA fraction on BinDel microdeletion risk detection sensitivity. Sensitivity estimates are indicated by grey bars (PCA95%)

**Suppl. Fig. S2.**
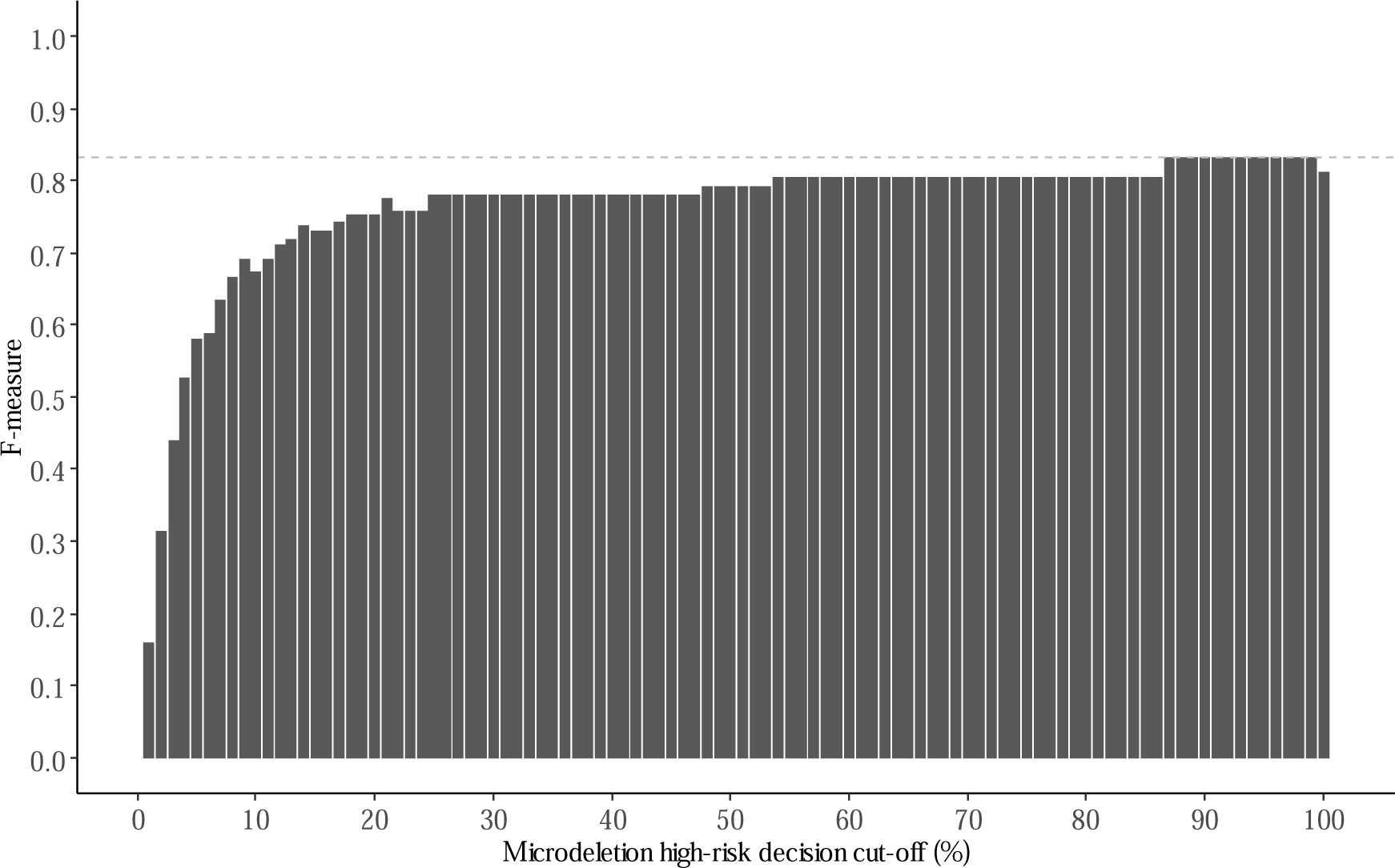
BinDel F-measure (the harmonic mean of positive predictive value and sensitivity, higher is better) on different microdeletion high-risk cut-off decision points with PCA95%.

**Suppl. Fig. S3.**
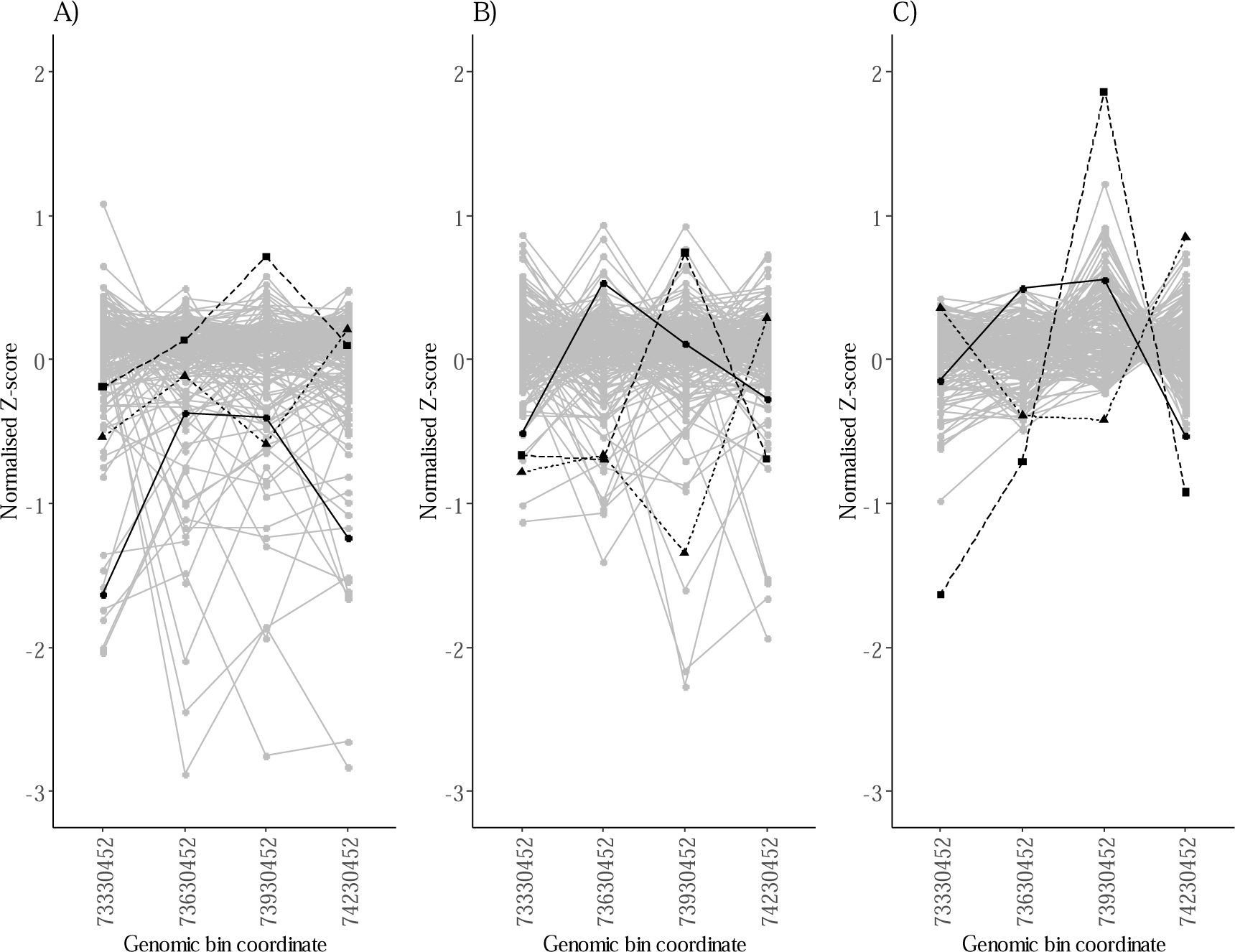
BinDel normalisation effects on genomic bin values in Williams-Beuren microdeletion region. (A) depicts normalised Z-scores (see **Methods**) without utilising the PCA normalisation method. (B) illustrates the impact of PCA95% normalisation on normalised Z-scores. (C) displays the combined effects of PCA95% and regional PCA (cumulative variance of 50%) normalised Z-scores. Each line in the Fig. represents a sample, where grey lines denote euploid reference samples, and black lines represent validation samples with fetal genomic microdeletion in the Williams-Beuren syndrome genomic region.

**Suppl. Table S1.** A priori chosen target microdeletion regions, their coordinates and bin sizes used in the analysis.

| **Coordinate**<br>**(chromosome:start-end)** | **Microdeletion region and database**<br>**used** | **Genomic**<br>**bin size**<br>**(kb)** |
| --- | --- | --- |
| 1:1-27 600 000 | 1p36 OMIM [24] | 300 |
| 1:10 001-12 780 116 | 1p36 DECIPHER [22] | 300 |
| 1:898 703-6 229 913 | 1p36 terminal region, ClinGen dosage ID: ISCA-37434 [22,23] | 300 |
| 3:192 600 000-198 295 559 | Chromosome 3q29 [24] | 300 |
| 4:1-4 500 000 | Wolf-Hirschhorn OMIM [24] | 300 |
| 4:337 779-2 009 235 | Wolf-Hirschhorn terminal region, ClinGen dosage ID: ISCA-37429 [22,23] | 300 |
| 4:1 567 470-2 108 509 | Wolf-Hirschhorn DECIPHER [22] | 100 |
| 5:10 001-12 533 192 | Cri-du-chat DECIPHER [22] | 300 |
| 7:72 700 001-77 900 000 | Williams-Beuren OMIM [24] | 300 |
| 7:73 330 452-74 728 334 | Williams-Beuren DECIPHER [22] | 300 |
| 8:116 700 000-126 300 000 | Langer-Giedion OMIM [24] | 300 |
| 11:114 600 000-135 086 622 | Jacobsen OMIM [24] | 300 |
| 15:22 677 345-28 134 728 | Prader-Willi/Angelman Type I. Start coordinate DECIPHER CNV syndrome, end coordinate ClinGen (dosage ID ISCA-37478) [22,23] | 300 |
| 15:23 374 765-28 134 728 | Prader-Willi/Angelman Type II. Start coordinate DECIPHER CNV syndrome, end coordinate ClinGen (dosage ID ISCA-37404) [22,23] | 300 |
| 17:16 869 758-20 318 836 | Smith-Magenis DECIPHER [22] | 300 |
| 17:30 780 079-31 936 302 | NF1 DECIPHER [22] | 200 |
| 20:80 106-1 311 812 | 20p13. Coordinates from a custom-ordered DNA Mix (lot #10560229) specification from SeraCare Life Sciences Inc with fetus DNA having a pathogenic loss of the terminal region of 20p13 and a pathogenic 3q29 duplication. | 300 |
| 22:18 924 718-20 299 685 | DiGeorge A-B, ClinGen dosage ID: ISCA-37433 [22,23] | 300 |
| 22:18 924 718-21 111 383 | DiGeorge A-D, ClinGen dosage ID: ISCA-37446 [22,23] | 300 |
| 22:20 377 696-21 111 383 | DiGeorge B/C-D, ClinGen dosage ID: ISCA-37516 [22,23] | 100 |

**Suppl. Table S2.**
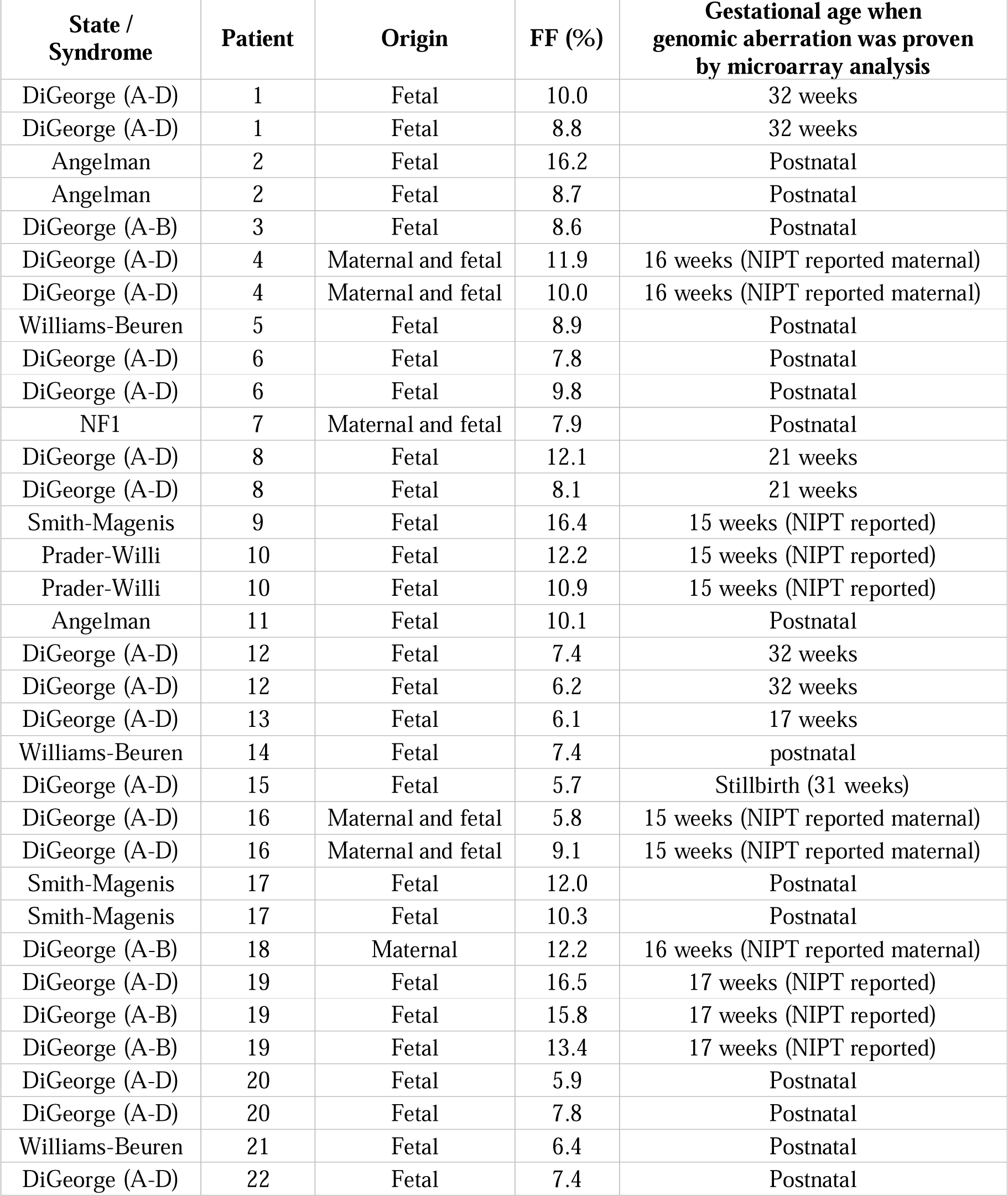
BinDel ‘blind’ validation sample set information. FF – Fetal fraction.

| **State / Syndrome** | **Patient** | **Origin** | **FF (%)** | **Gestational age when genomic aberration was proven by microarray analysis** |
| --- | --- | --- | --- | --- |
| DiGeorge (A-D) | 1 | Fetal | 10.0 | 32 weeks |
| DiGeorge (A-D) | 1 | Fetal | 8.8 | 32 weeks |
| Angelman | 2 | Fetal | 16.2 | Postnatal |
| Angelman | 2 | Fetal | 8.7 | Postnatal |
| DiGeorge (A-B) | 3 | Fetal | 8.6 | Postnatal |
| DiGeorge (A-D) | 4 | Maternal and fetal | 11.9 | 16 weeks (NIPT reported maternal) |
| DiGeorge (A-D) | 4 | Maternal and fetal | 10.0 | 16 weeks (NIPT reported maternal) |
| Williams-Beuren | 5 | Fetal | 8.9 | Postnatal |
| DiGeorge (A-D) | 6 | Fetal | 7.8 | Postnatal |
| DiGeorge (A-D) | 6 | Fetal | 9.8 | Postnatal |
| NF1 | 7 | Maternal and fetal | 7.9 | Postnatal |
| DiGeorge (A-D) | 8 | Fetal | 12.1 | 21 weeks |
| DiGeorge (A-D) | 8 | Fetal | 8.1 | 21 weeks |
| Smith-Magenis | 9 | Fetal | 16.4 | 15 weeks (NIPT reported) |
| Prader-Willi | 10 | Fetal | 12.2 | 15 weeks (NIPT reported) |
| Prader-Willi | 10 | Fetal | 10.9 | 15 weeks (NIPT reported) |
| Angelman | 11 | Fetal | 10.1 | Postnatal |
| DiGeorge (A-D) | 12 | Fetal | 7.4 | 32 weeks |
| DiGeorge (A-D) | 12 | Fetal | 6.2 | 32 weeks |
| DiGeorge (A-D) | 13 | Fetal | 6.1 | 17 weeks |
| Williams-Beuren | 14 | Fetal | 7.4 | postnatal |
| DiGeorge (A-D) | 15 | Fetal | 5.7 | Stillbirth (31 weeks) |
| DiGeorge (A-D) | 16 | Maternal and fetal | 5.8 | 15 weeks (NIPT reported maternal) |
| DiGeorge (A-D) | 16 | Maternal and fetal | 9.1 | 15 weeks (NIPT reported maternal) |
| Smith-Magenis | 17 | Fetal | 12.0 | Postnatal |
| Smith-Magenis | 17 | Fetal | 10.3 | Postnatal |
| DiGeorge (A-B) | 18 | Maternal | 12.2 | 16 weeks (NIPT reported maternal) |
| DiGeorge (A-D) | 19 | Fetal | 16.5 | 17 weeks (NIPT reported) |
| DiGeorge (A-B) | 19 | Fetal | 15.8 | 17 weeks (NIPT reported) |
| DiGeorge (A-B) | 19 | Fetal | 13.4 | 17 weeks (NIPT reported) |
| DiGeorge (A-D) | 20 | Fetal | 5.9 | Postnatal |
| DiGeorge (A-D) | 20 | Fetal | 7.8 | Postnatal |
| Williams-Beuren | 21 | Fetal | 6.4 | Postnatal |
| DiGeorge (A-D) | 22 | Fetal | 7.4 | Postnatal |

**Suppl. Table S3.**
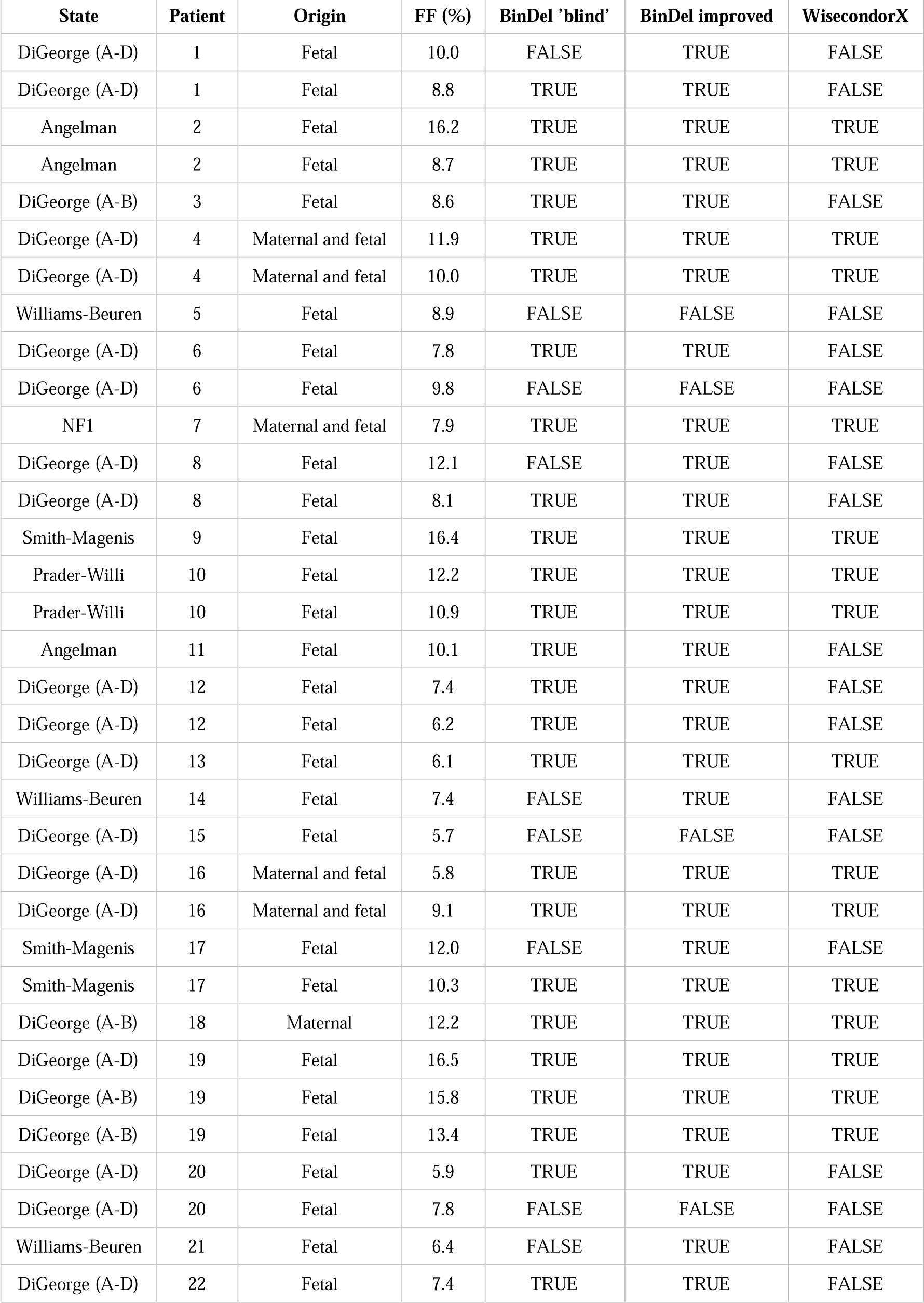
BinDel and WisecondorX microdeletion calls on the validation sample set. FF – Fetal fraction.

